# Specificity of a polygenic score for aggressive prostate cancer

**DOI:** 10.1101/2025.01.23.25321041

**Authors:** Anna M Dornisch, George J Xu, Roshan Karunamuni, Charles A Brunette, Morgan E Danowski, Craig C Teerlink, J. Michael Gaziano, Isla P Garraway, Richard L Hauger, Adam S Kibel, Julie A Lynch, Kara N Maxwell, Brent S Rose, Ole A Andreassen, Anders M Dale, Jenny L Donovan, Freddie Hamdy, Athene Lane, Ian G Mills, Richard M Martin, David E Neal, Emma L Turner, Alicja Wolk, PRACTICAL Consortium, VA Million Veteran Program, Jason L Vassy, Tyler Seibert

## Abstract

To address the concern that polygenic hazard scores for prostate cancer (PCa) might not distinguish between indolent and aggressive disease, we performed *case-only* analyses using a 601-variant polygenic score (PHS601). We hypothesized that among men who eventually developed PCa, those with higher PHS were more likely to develop aggressive disease.

We analyzed genetic and phenotypic data from a diverse, national cohort of men diagnosed with PCa (Million Veteran Program, n = 69,901, 6413 metastatic). We used Cox proportional hazards models to examine the association of PHS601 with both age at onset of metastatic PCa (birth-to-met) and time from localized to metastatic diagnosis (localized-to-met). We performed an external validation of the case-only birth-to-met analysis in two population-based cohorts from the PRACTICAL Consortium.

PHS601 was associated with both birth-to-met and localized-to-met within MVP. The HRs for men in the highest quintile of PHS601 vs. those in the lowest quintile (HR_80/20_) were 1.74 [1.65-1.84] and 1.41 [1.31-1.41] for birth-to-met and localized-to-met, respectively. These findings were validated in two external cohorts (COSM and ProtecT). PHS601 was also associated with earlier development of metastasis. Men with high PHS601 are diagnosed with prostate cancer at a younger age and are more likely to develop metastasis.

## Manuscript

Metastatic prostate cancer is a major public health challenge that is both worsening in frequency yet potentially preventable^1^. Given limited resources and the known harms of screening, efforts at early detection of prostate cancer should focus on those at highest risk of developing metastatic disease. Although screening guidelines vary, common tenets include shared decision making and individual risk assessment based on family history and self-reported race^2,3^. Such risk assessment is subjective and inconsistent.

Genetic risk stratification has emerged as a promising means to objectively and accurately assess lifetime risk of prostate cancer^4^. Polygenic hazard scores (PHS) are strongly associated with age at diagnosis of any prostate cancer, as well as the lifetime risk of metastatic and fatal prostate cancer^5–7^. However, because these scores are associated with any prostate cancer, there is concern that screening men with high polygenic risk could increase overdiagnosis of indolent cancers. To address this, we performed case-only analyses using a 601-variant polygenic score (PHS601) that is also used in an ongoing national randomized controlled trial of genomics-informed, precision prostate cancer screening (ProGRESS; NCT05926102)^8^. We hypothesized that among men who eventually developed prostate cancer, those with higher PHS were more likely to develop aggressive disease.

We analyzed genetic and phenotypic data from a diverse, national cohort of men diagnosed with prostate cancer (Million Veteran Program; n=69,901; 6,413 metastatic). We used univariable Cox proportional hazards models to test PHS601 for association with age at onset of metastatic prostate cancer (birth-to-met analysis) and time between diagnoses of localized and metastatic prostate cancer (localized-to-met). We also conducted multivariable Cox analyses to test PHS601 for association with localized-to-met while accounting for stage and age at diagnosis. For both endpoints, we considered definitive treatment for clinically localized disease to alter the natural disease progression^9^. Participants who were treated for clinically localized disease and still later experienced metastasis were considered to have been treated too late (or inadequately). Thus, onset of metastasis was defined as time of first treatment or time of diagnosis of metastasis, whichever occurred first. Participants who never experienced metastasis were right censored at time of first treatment (if applicable) or time of last follow-up/death. The Cox model estimates were used to generate hazard ratios (HR) comparing men in various percentile groups (e.g., top 20^th^ vs. bottom 20^th^, obtaining these thresholds within men of European genetic ancestry for reference)^5^. We used 100-fold bootstrapping to estimate 95% confidence intervals and generated cause-specific cumulative incidence curves for each endpoint as previously described^7^. Since PHS601 was originally developed in the MVP dataset (albeit with any prostate cancer— not metastasis—as the outcome), we performed an external validation of the case-only, birth-to-met analysis in two population-based cohorts from the PRACTICAL Consortium (Cohort of Swedish Men [COSM, n=2,163], Prostate Testing for Cancer and Treatment [ProtecT, n=1,583]). Given the much smaller size of these cohorts and that metastatic outcome is not available in these datasets, we used birth-to-clinically-significant prostate cancer (NCCN intermediate or higher risk) as the outcome to evaluate consistency of association with more aggressive disease.

PHS601 was associated with both birth-to-met and localized-to-met within MVP. The HRs for men in the highest quintile of PHS601 versus those in the lowest quintile (HR_80/20_) were 1.74 [1.65-1.84] and 1.41 [1.3-1.41] for birth-to-met and localized-to-met, respectively. Within the external validation PRACTICAL cohorts, PHS601 was associated with the development of clinically significant prostate cancer. HR_80/20_ within the COSM and ProtecT data were 1.33 [1.15-1.52] and 1.64 [1.36-2.04], respectively. Cumulative incidence curves are shown in Figure 1. For comparison, a prior study found that a 400-variant polygenic score was associated with ~5% increased odds of aggressive localized or metastatic disease per standard deviation of the score^10^.

**Figure 1:**
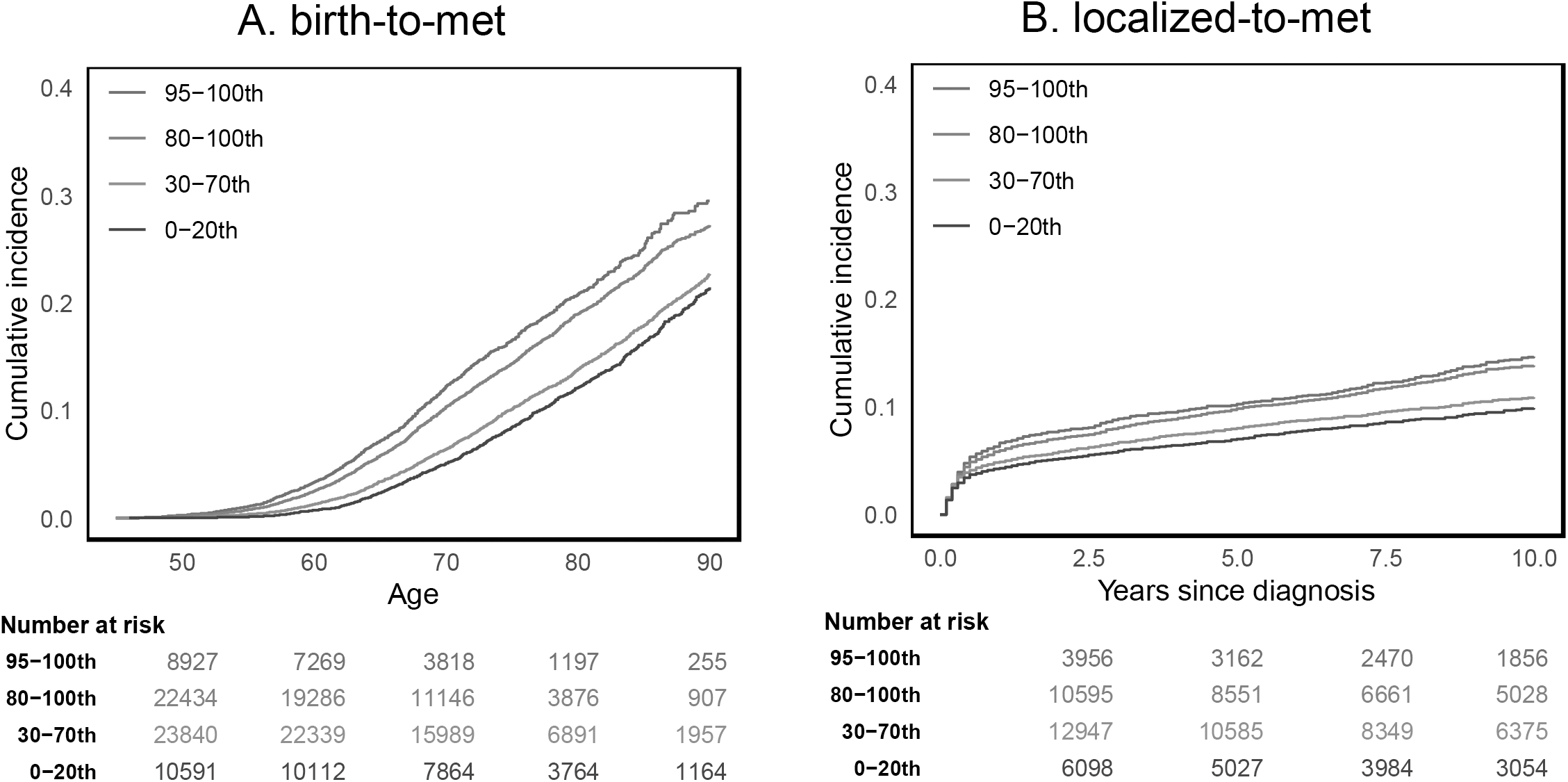
Cumulative incidence of metastatic prostate cancer in MVP by PHS601 strata. Cumulative incidence within MVP, stratified by 601-variant polygenic hazard score (PHS601), for (A) birth-to-met and (B) univariable localized-to-met. Colors represent various strata of PHS601: 0-20th, 30-70th, 80-100th, and 95-100^th^ percentiles (cutoffs defined within men of European ancestry).

Among men eventually diagnosed with prostate cancer, those with higher polygenic risk were more likely to develop metastatic disease. Compared to participants with low PHS601 (bottom quintile), those above the 95^th^ and 80^th^ percentiles had an absolute increase of 8.1% [6.5-9.7%] and 5.9% [4.8-7.1%] in incidence of metastasis by age 75, respectively. They reached an incidence of metastasis equivalent to that of 75-year-old participants with low PHS601 8.3 [7.8-9.1] and 7.0 [6.3-7.5] years younger, respectively.

When accounting for PSA, stage, and age at diagnosis, HR_80/20_ for localized-to-met was 1.14 [1.04-1.27], which is significant but not as strong as available biomarkers based on tumor gene expression or pathology artificial intelligence. Germline polygenic scores appear most useful to stratify risk for screening and early detection.

To leverage greater statistical power for model training, PHS601 was developed with age at diagnosis of any prostate cancer as the time-to-event outcome. Concerns arise that risk stratification with PHS601 might lead to increased overdiagnosis of indolent disease. However, without any modification or tuning of parameters, we found PHS601 is moderately specific to risk of metastatic disease despite not being explicitly trained for this outcome. This may be partially because men who develop prostate cancer younger might also be more likely to develop metastatic prostate cancer at an earlier age. Thus, time-to-event models trained on a higher frequency event (any prostate cancer), may provide stable parameter estimates that can be successfully applied to lower frequency events (metastasis) without re-training if the latter event is dependent on the former.

As genetic risk scores like PHS601 can be performed on a saliva sample at *any time* during a person’s life, they provide the earliest information about age-specific risk of developing aggressive prostate cancer^4^. These scores might be useful to support clinical decisions not only about *whom* to screen but also at what age.

Despite using methods to limit the impact of potential overfitting, HR values could be somewhat inflated in MVP because PHS601 was developed using that data. We have addressed this by demonstrating comparable effect sizes for the birth-to-met analysis in two external validation datasets, with the caveat that we had to use a proxy outcome in the validation sets.

Men with high PHS601 are diagnosed with prostate cancer at younger age and are more likely to develop metastasis.

## Data Availability

All data produced in the present study are available upon reasonable request to the authors

## ACKNOWLEDGEMENT AND FUNDING STATEMENT

**Acknowledgements**

This publication does not represent the views of the Department of Veterans Affairs or the United States Government.

## VA Million Veteran Program Core Acknowledgement for Publications. MVP Program Office

Sumitra Muralidhar, Ph.D., Program Director, US Department of Veterans Affairs, 810 Vermont Avenue NW, Washington, DC 20420; Jennifer Moser, Ph.D., Associate Director, Scientific Programs, US Department of Veterans Affairs, 810 Vermont Avenue NW, Washington, DC 20420; Jennifer E. Deen, B.S., Associate Director, Cohort & Public Relations, US Department of Veterans Affairs, 810 Vermont Avenue NW, Washington, DC 20420. **MVP Executive Committee:** Co-Chair: Philip S. Tsao, Ph.D., VA Palo Alto Health Care System, 3801 Miranda Avenue, Palo Alto, CA 94304; Co-Chair: Sumitra Muralidhar, Ph.D. US Department of Veterans Affairs, 810 Vermont Avenue NW, Washington, DC 20420; J. Michael Gaziano, M.D., M.P.H., A Boston Healthcare System, 150 S. Huntington Avenue, Boston, MA 02130; Elizabeth Hauser, Ph.D., Durham VA Medical Center, 508 Fulton Street, Durham, NC 27705;, Amy Kilbourne, Ph.D., M.P.H., VA HSR&D, 2215 Fuller Road, Ann Arbor, MI 48105; Michael Matheny, M.D., M.S., M.P.H., VA Tennessee Valley Healthcare System, 1310 24^th^ Ave. South, Nashville, TN 37212;, Dave Oslin, M.D., Philadelphia VA Medical Center, 3900 Woodland Avenue, Philadelphia, PA 19104;, Deepak Voora, MD Durham VA Medical Center, 508 Fulton Street, Durham, NC 27705. **MVP Co-Principal Investigators:** J. Michael Gaziano, M.D., M.P.H., VA Boston Healthcare System, 150 S. Huntington Avenue, Boston, MA 02130; Philip S. Tsao, Ph.D.. VA Palo Alto Health Care System, 3801 Miranda Avenue, Palo Alto, CA 94304. **MVP Core Operations:** Jessica V. Brewer, M.P.H., Director, MVP Recruitment & Enrollment VA Boston Healthcare System, 150 S. Huntington Avenue, Boston, MA 02130; Mary T. Brophy M.D., M.P.H., Director, VA Central Biorepository, VA Boston Healthcare System, 150 S. Huntington Avenue, Boston, MA 02130; Kelly Cho, M.P.H, Ph.D., Director, MVP Phenomics Data Core, VA Boston Healthcare System, 150 S. Huntington Avenue, Boston, MA 02130; Lori Churby, B.S., Director, MVP Regulatory Affairs VA Palo Alto Health Care System, 3801 Miranda Avenue, Palo Alto, CA 94304; Scott L. DuVall, Ph.D., Director, VA Informatics and Computing Infrastructure (VINCI) VA Salt Lake City Health Care System, 500 Foothill Drive, Salt Lake City, UT 84148; Saiju Pyarajan Ph.D., Director, Data and Computational Sciences VA Boston Healthcare System, 150 S. Huntington Avenue, Boston, MA 02130; Robert Ringer, Pharm.D., Director, VA Albuquerque Central Biorepository, New Mexico VA Health Care System, 1501 San Pedro Drive SE, Albuquerque, NM 87108; Luis E. Selva, Ph.D., Executive Director, MVP Biorepositories, VA Boston Healthcare System, 150 S. Huntington Avenue, Boston, MA 02130; Shahpoor (Alex) Shayan, M.S., Director, MVP Recruitment and Enrollment Informatics, VA Boston Healthcare System, 150 S. Huntington Avenue, Boston, MA 02130; Brady Stephens, M.S., Principal Investigator, MVP Information Center, Canandaigua VA Medical Center, 400 Fort Hill Avenue, Canandaigua, NY 14424; Stacey B. Whitbourne, Ph.D., Director, MVP Cohort Management VA Boston Healthcare System, 150 S. Huntington Avenue, Boston, MA 02130. **MVP Publications and Presentations Committee:** Co-Chair: Themistocles L. Assimes, M.D., Ph. D, VA Palo Alto Health Care System, 3801 Miranda Avenue, Palo Alto, CA 94304; Co-Chair: Adriana Hung, M.D.; M.P.H, VA Tennessee Valley Healthcare System, 1310 24^th^ Ave. South, Nashville, TN 37212; Co-Chair: Henry Kranzler, M.D., Philadelphia VA Medical Center, 3900 Woodland Avenue, Philadelphia, PA 19104.

## DISCLOSURES

AMD reports honoraria from Conquer Cancer. JAL reports grants from Alnylam Pharmaceuticals, Inc., Astellas Pharma, Inc., AstraZeneca Pharmaceuticals LP, Biodesix, Inc, Celgene Corporation, Cerner Enviza, GSK PLC, IQVIA Inc., Janssen Pharmaceuticals, Inc., Novartis International AG, Parexel International Corporation through the University of Utah or Western Institute for Veteran Research outside the submitted work. ASK reports fundings (for work unrelated to this publication) from Janssen, Pfizer, Profound, Bristol Myers Squibb, and Merck. AMD reports honoraria from Conquer Cancer. AMD is a founder of and holds equity interest in CorTechs Labs and serves on its scientific advisory board. He is also a member of the Scientific Advisory Board of Healthlytix and receives research funding from General Electric Healthcare (GEHC).TMS reports honoraria from Varian Medical Systems, WebMD, GE Healthcare, and Janssen; he has an equity interest in CorTechs Labs, Inc. and serves on its Scientific Advisory Board; he receives research funding from GE Healthcare through the University of California San Diego. These companies might potentially benefit from the research results. The terms of this arrangement have been reviewed and approved by the University of California San Diego in accordance with its conflict-of-interest policies. The other authors have no disclosures.

TABLES AND FIGURES (2 total, others can go to supplement):

**Table 1.** Association of PHS601 with birth-to-met and localized-to-met in MVP and PRACTICAL cohorts. Data are HR (95% CI) for birth-to-met in MVP, COSM, and ProtecT as well as HR (95% CI) for localized-to-met in MVP in both univariable analysis and multivariable analyses. PHS601 remains significantly associated with localized-to-met when accounting for age and AJCC stage group at diagnosis. ^a^In COSM and ProtecT, clinically significant prostate cancer was defined as Gleason score ≥7, PSA ≥10 ng/mL, T3-T4 stage, nodal metastases, or distant metastases

| Model | Birth-to-met |  |  | Localized-to-met |  |
| --- | --- | --- | --- | --- | --- |
| Cohort | MVP | COSM<sup>a</sup> | ProtecT<sup>a</sup> | MVP - univariable | MVP-multivariable |
| **PHS601** |  |  |  |  |  |
| HR<sub>80/20</sub> | 1.74<br>[1.64,1.84] | 1.33<br>[1.15,1.52] | 1.64<br>[1.36,2.04] | 1.41<br>[1.31,1.51] | 1.20<br>[1.08,1.31] |
| HR<sub>80/50</sub> | 1.35<br>[1.32,1.40] | 1.16<br>[1.08,1.24] | 1.29<br>[1.17,1.44] | 1.20<br>[1.16,1.25] | 1.10<br>[1.04,1.16] |
| HR<sub>20/50</sub> | 0.78<br>[0.76,0.80] | 0.87<br>[0.82,0.94] | 0.79<br>[0.70,0.86] | 0.86<br>[0.83,0.89] | 0.92<br>[0.88,0.96] |
| HR<sub>95/50</sub> | 1.53<br>[1.47,1.60] | 1.25<br>[1.12,1.39] | 1.47<br>[1.27,1.75] | 1.30<br>[1.23,1.37] | 1.15<br>[1.06,1.24] |
| **Age (years)\*** |  |  |  |  | 1.06<br>[1.01,1.12] |
| **Stage** |  |  |  |  |  |
| II vs. I |  |  |  |  | 2.97<br>[2.62,3.33] |
| III vs. I |  |  |  |  | 7.66<br>[6.60,8.82] |

\* Hazard ratio for Age at diagnosis is for every 10-year increase

## REFERENCES

1. James, N. D. et al. The Lancet Commission on prostate cancer: planning for the surge in cases. The Lancet (2024) doi:10.1016/S0140-6736(24)00651-2.

2. Moses, K. A. et al. NCCN Guidelines® Insights: Prostate Cancer Early Detection, Version 1.2023: Featured Updates to the NCCN Guidelines. J. Natl. Compr. Canc. Netw. 21, 236–246 (2023).

3. Cornford, P. et al. EAU-EANM-ESTRO-ESUR-ISUP-SIOG Guidelines on Prostate Cancer—2024 Update. Part I: Screening, Diagnosis, and Local Treatment with Curative Intent. Eur. Urol. (2024) doi:10.1016/j.eururo.2024.03.027.

4. Seibert, T. M. et al. Genetic Risk Prediction for Prostate Cancer: Implications for Early Detection and Prevention. Eur. Urol. (2023) doi:10.1016/j.eururo.2022.12.021.

5. Seibert, T. M. et al. Polygenic hazard score to guide screening for aggressive prostate cancer: development and validation in large scale cohorts. BMJ 360, j5757 (2018).

6. Huynh-Le, M.-P. et al. Polygenic hazard score is associated with prostate cancer in multi-ethnic populations. Nat. Commun. 12, 1236 (2021).

7. Pagadala, M. S. et al. Polygenic risk of any, metastatic, and fatal prostate cancer in the Million Veteran Program. JNCI J. Natl. Cancer Inst. 115, 190–199 (2023).

8. Vassy, J. L. et al. From a genomic risk model to clinical trial implementation in a learning health system: the ProGRESS Study. 2024.11.03.24316516 Preprint at 10.1101/2024.11.03.24316516 (2024).

9. Hamdy, F. C. et al. Fifteen-Year Outcomes after Monitoring, Surgery, or Radiotherapy for Prostate Cancer. N. Engl. J. Med. 0, null (2023).

10. Wang, A. et al. Characterizing prostate cancer risk through multi-ancestry genome-wide discovery of 187 novel risk variants. Nat. Genet. 1–10 (2023) doi:10.1038/s41588-023-01534-4.

