## Supplementary Table 1-3 for "Specificity of a polygenic score for aggressive prostate cancer"

**Supplementary Table 1. MVP participant breakdown**

|  | | Birth-to-met | Localized-to-met  (univariable) | Localized-to-met  (multivariable) |
| --- | --- | --- | --- | --- |
| Number of cases | | 69,901 | 65,006 | 27,840 |
| Number of metastases | | 8,191 | 5,588 | 3,094 |
| Age at enrollment in MVP (years) | | 70.0  [25.1,103.7] | 70.0  [25.1,103.7] | 68.2 [33.9,103.7] |
| Age at diagnosis | | 67.4  [24.2,104.9] | 67.2  [24.2,104.9] | 65.4  [31.9,95.9] |
| Age at metastasis | | 73.0  [38, 104.5] | 73.5  [38,104.5] | 72.2  [38,99.8] |
| Age at first treatment | | 68.9  [30.4,102.3] | 68.7  [30.4,102.3] | 66.7  [36.8,99.5] |
| Age at last follow-up | | 77.9  [33.1,110.2] | 78.0  [33.1,110.2] | 76.6  [44.6,105] |
| Stage at diagnosis (n) | I | 5,756 | 5,568 | 5,568 |
|  | II | 20,235 | 19,400 | 19,400 |
|  | III | 3,044 | 2,872 | 2,872 |
|  | IV | 2,085 | 0 | 0 |

* Continuous variables are reported as median [min,max]

**Supplementary Table 2. PRACTICAL participant breakdown**

|  | ProtecT | COSM |
| --- | --- | --- |
| Number of cases | 1,583 | 2,163 |
| Number of clinically significant cases | 628 | 1,403 |
| Age at diagnosis (years) | 63.4  [45,71.8] | 70.0  [49.6,92] |
| Age at clinically significant diagnosis | 64.3  [49.3,71.8] | 72.4  [49.6,92] |
| Age at last follow-up | 63.4  [45,71.8] | 77.9  [58.1,96.9] |

* Continuous variables are reported as median [min,max]

**Supplementary Table 3. Multivariable MVP analysis for localized-to-met with PHS601, Age, PSA and Grade Group at diagnosis (n=29,510).**

|  | | Localized-to-met  MVP-multivariable |
| --- | --- | --- |
| PHS601 | HR_80/20_ | 1.05  [0.95,1.17] |
|  | HR_80/50_ | 1.03  [0.97,1.09] |
|  | HR_20/50_ | 0.98  [0.93,1.02] |
|  | HR_95/50_ | 1.04  [0.96,1.13] |
| Age (years)* | | 0.86  [0.80,0.92] |
| PSA (ng/mL)* | | 1.04  [1.03,1.11] |
| Grade Group | 2 vs. 1 | 2.00  [1.80,2.23] |
|  | 3 vs. 1 | 3.60  [3.26,4.03] |
|  | 4 vs. 1 | 5.83  [5.04,6.46] |
|  | 5 vs. 1 | 9.85  [8.59,10.99] |

**Hazard ratio for Age and PSA at diagnosis are reported for every 10-unit increase in the variable*
